# Prevalence of Mild and Severe Cognitive Impairment in World Trade Center Exposed Fire Department of the City of New York (FDNY) and General Emergency Responders

**DOI:** 10.1101/2024.08.04.24311457

**Authors:** Frank D. Mann, Alexandra K. Mueller, Rachel Zeig-Owens, Jaeun Choi, David J. Prezant, Melissa M. Carr, Alicia M. Fels, Christina M. Hennington, Megan P. Armstrong, Alissa Barber, Ashley E. Fontana, Cassandra H. Kroll, Kevin Chow, Onix A. Melendez, Abigail J. Smith, Benjamin J. Luft, Charles B. Hall, Sean A. P. Clouston

## Abstract

**Background:** The emergency personnel who responded to the World Trade Center (WTC) attacks endured severe occupational exposures, yet the prevalence of cognitive impairment remains unknown among WTC-exposed-FDNY-responders. The present study screened for mild and severe cognitive impairment in WTC-exposed FDNY responders using objective tests, compared prevalence rates to a cohort of non-FDNY WTC-exposed responders, and descriptively to meta-analytic estimates of MCI from global, community, and clinical populations.

**Methods:** A sample of WTC-exposed-FDNY responders (n = 343) was recruited to complete an extensive battery of cognitive, psychological, and physical tests. The prevalences of domain-specific impairments were estimated based on the results of norm-referenced tests, and the Montreal Cognitive Assessment (MoCA), Jak/Bondi criteria, Petersen criteria, and the National Institute on Aging and Alzheimer’s Association (NIA-AA) criteria were used to diagnose MCI. NIA-AA criteria were also used to diagnose severe cognitive impairment. Generalized linear models were used to compare prevalence estimates of cognitive impairment to a large sample of WTC-exposed-non-FDNY responders from the General Responder Cohort (GRC; n = 7102) who completed the MoCA during a similar time frame.

**Result:** Among FDNY responders under 65 years, the unadjusted prevalence of MCI varied from 52.57% to 71.37% depending on the operational definition of MCI, apart from using a conservative cut-off applied to MoCA total scores (18 < MoCA < 23), which yielded a markedly lower crude prevalence (24.31%) compared to alternative criteria. The prevalence of MCI was higher among WTC-exposed-FDNY-responders, compared to WTC-exposed-non-FDNY-GRC-responders (adjusted *RR* = 1.53, *95% C.I*. = [1.24, 1.88], *p* < .001) and meta-analytic estimates from different global, community, and clinical populations. Following NIA-AA diagnostic guidelines, 4.96% of WTC-exposed-FDNY-responders met the criteria for severe impairments (95% CI = [2.91% to 7.82%]), a prevalence that remained largely unchanged after excluding responders over the age of 65 years.

**Discussion:** There is a high prevalence of mild and severe cognitive impairment among WTC-responders highlighting the putative role of occupational/environmental and disaster-related exposures in the etiology of accelerated cognitive decline.

## Prevalence of Mild and Severe Cognitive Impairment in World Trade Center Exposed Fire Department of the City of New York (FDNY) and General Emergency Responders

During and after the 9/11/2001 (9/11) attacks on the World Trade Center (WTC), members of the Fire Department of the City of New York (FDNY) and other emergency personnel inhaled hazardous particulate matter from the dust cloud that was expelled from the combustion of many thousands of pounds of jet fuel and the collapse of the Twin Towers^1^. While an emerging body of research has clarified the extent of cognitive impairments in police and volunteers who responded to the 9/11 attacks^2,3^, FDNY responders are different in their training and in the natures of their exposures, compared with other emergency personnel. FDNY responders, including firefighters and emergency medical service (EMS) providers, were among the first to arrive at the WTC-site and endured prolonged exposures digging through the rubble to search for survivors. Crucially, the neurocognitive impact of exposures that might be unique to FDNY responders remains unknown, and no study to date has systematically screened for cognitive impairments in this well characterized occupational cohort. Consequently, for WTC-exposed FDNY responders the prevalences of mild and severe cognitive impairments remain unknown. Previous research has reported subjective cognitive concerns among WTC-exposed firefighters^4^. Yet, a large multi-cohort study found that WTC-exposed FDNY firefighters reported fewer cognitive concerns than firefighters from Chicago, Philadelphia, and San Francisco who were not exposed to the WTC attacks^5^, rendering unclear the clinical significance of self-reported cognitive concerns and correspondence to objectively measured deficits in cognitive function.

The prevalence of cognitive impairment increases sharply with age, including both mild cognitive impairment (MCI ) and more severe impairment possibly indicative of a major neurocognitive disorder (NCD), the term now used in the DSM-V^6^ to replace that previously referred to as dementia in the DSM-IV-TR^7^. MCI is fairly common among middle-aged adults, with meta-analytic estimates of prevalence ranging from 10% to 25% across community samples of adults 50 years and older^8^. MCI is also common in clinical populations, evinced by meta-analyses of residents in nursing homes^9,10^, and patients with Parkinson’s disease^11^, clinical hypertension^12^, scarpenia^13^, ischemic and hemorrhagic stroke^14^, and breast cancer during chemotherapy^15^. On the other hand, major NCDs from any cause is very uncommon before the age of 65, with estimates ranging from 0.01% to 0.50% across different meta-analyses and large population-based studies^16–20^. Consequently, if WTC-related occupational exposures are unrelated to cognitive impairment, then the prevalence rates of mild and severe cognitive impairment among the WTC-exposed should be similar, if not lower, to those reported in these studies, particularly among those under 65 years old.

The present study fills several gaps in knowledge. First, the prevalence of MCI is estimated and compared using the same diagnostic criteria in different cohorts of WTC-exposed responders, as the potential for severe pathogenic exposures is hypothesized to be more common among FDNY responders compared to responders from the General Responder Cohort (GRC), who were not members of the FDNY on 9/11/2001 and consist mostly of law enforcement officers, construction and communications workers, and civilian volunteers, henceforth called GRC responders. Our recent work highlights the central role of long-term exposures to fine particulate matter in relation to cognitive impairment in GRC responders^3^, and while these responders were exposed to known toxins, they may have been exposed to different types of particulate matter and for different lengths of time as compared to FDNY responders. Therefore, the primary objective of the present study was to understand whether FDNY and GRC responders differ in the prevalence of MCI. Secondary objectives include estimating the prevalence of severe cognitive impairment indicative of a major NCD, as well as the prevalence of domain specific impairments based on objective norm-referenced tests, and the cognitive, psychological, and physical functional correlates of cognitive impairment in FDNY responders. Finally, to assess the potential impact of different operational definitions on prevalence rates, the prevalence of MCI was estimated among WTC-exposed FDNY responders using different diagnostic criteria.

It is critical and timely to study the prevalence of MCI and possible major NCD in FDNY responders for at least three reasons. First, older members of the cohort are now approaching the age at which cognitive impairment becomes more common. Second, other studies have documented mild and severe cognitive impairment in GRC responders before the age of 65 years^21^. Third, the unique exposures endured by members of the FDNY could have resulted in an even greater impact on cognition. Knowing the prevalences of mild and severe cognitive impairment will enable future research to identify exposure-response pathways and biomarker characterization. Moreover, by measuring cognitive and physical functional deficits in FDNY responders using a wide range of objective tests, the present study extends our previous research by more thoroughly characterizing the domain-specific deficits of cognitively impaired WTC responders.

## Method

### Sample

#### Source population

The population provided written informed consent and included firefighters and EMS providers present at the WTC disaster site for at least one day between 9/11/2001 and 7/24/2002. We restricted the source population to members born between 1/1/1952 and 12/31/1981 who were alive at the time of recruitment. Males were further restricted to those residing in Long Island (Suffolk or Nassau counties) where study activities took place. Due to fewer females in the FDNY population, the recruitment pool of female participants was expanded to include those residing in Long Island, Brooklyn, and Queens. The source population included 4,198 potential participants (Table S1).

WTC-exposed FDNY responders must: 1) speak English fluently, 2) complete a written examination, 3) be a U.S. citizen at the time of appointment, 4) have completed high school or its equivalent, or be honorably discharged from the military, 5) pass medical, social, and psychological screening tests, and 6) undergo rigorous training. All WTC-exposed responders continue to receive healthcare and monitoring for WTC-related illnesses pursuant to the James Zadroga 9/11 Health and Compensation Act. As such, these criteria applied to FDNY source population.

#### Participant Recruitment

FDNY research team members sent weekly recruitment letters to randomly selected surviving FDNY responders from the source population. While individuals were randomly selected to receive a letter, participants voluntarily chose whether to respond and enroll in the study, introducing the potential for self-selection bias which can lead to a non-representative sample.

Examination of early enrollment found those enrolling had more subjective cognitive concerns \than the source population, as measured by the cognitive function instrument (CFI)^22^. To account for this, we took a targeted-outreach approach and restricted further mailings to those with no prior reports of subjective cognitive concerns. This targeted approach provided a study sample that is representative of the source population (Table S1).

Letters explained the purpose of the research and a point of contact for enrolling in the study. Additionally, the letter stated that the member may be contacted by research staff if they had previously consented to such contact. Follow-up letters were sent to anyone who did not contact study coordinators within one month of the original letter. Meanwhile, research staff at Stony Brook University began telephone calls to all prospective participants who had 1) received a letter in the first round of mailing, 2) agreed to allow non-FDNY researchers to contact them about their participation in the WTC events (43.3% of the population), and 3) had a phone number listed (1.5% did not). Eligible participants received phone calls the following weeks and left a message, if needed. During the initial contact call (either incoming or staff initiated), research staff would determine if the prospective participant was eligible based on inclusion and exclusion criteria.

To be eligible for the study, participants had to be willing to travel to one of the research sites to complete the study and had to consent to undergo a blood draw by a trained phlebotomist. Participants were excluded from the study if they already had a known neurological disorder that could cause cognitive impairment like brain cancer, stroke, or a traumatic head injury. All protocols were approved by the Institutional Review Board at Stony Brook University (CORIHS IRB#2021-00295), and participants provided informed oral and written consent. Only after obtaining consent, research staff proceeded with data collection. To maintain the confidentiality of case status for participants, all recruitment and data collection was done at research sites maintained by Stony Brook University under a certificate of confidentiality. Data collection was conducted from 11/30/2021 to 12/15/2023.

#### Comparison Cohort

To better contextualize levels of MCI in this trauma-exposed occupational cohort, estimates of prevalence from FDNY responders were compared to a large cohort of GRC responders who were predominately law enforcement officers, constructions workers, medical personnel, and other volunteer responders (*n* = 7102). Drawn from a large ongoing prospective study of cognitive impairment, GRC responders are eligible to participate in free annual monitoring visits at health clinics that are in the same buildings as the FDNY research sites described above. Inclusion criteria for the comparison cohort include (1) having completed a MoCA during a similar window of data collection as the FDNY cohort (11/30/2021 to 4/28/2023) and (2) being 43 to 71 years old at the time of data collection to match the age range of enrolled FDNY participants. Exclusion criteria for both the FDNY and the comparison GRC cohort included having a known neurological disorder that could cause cognitive impairment. Previous work has demonstrated that this study sample is broadly representative of the source population from which it was drawn^2^.

### Measures

#### Mild Cognitive Impairment (MCI)

The *Montreal Cognitive Assessment (MoCA),* a standard clinical assessment that integrates information from twelve short-form versions of commonly used tests of executive function, memory, and visual-spatial capability, was administered by trained study personnel and scored using a validated algorithm that was designed to be sensitive to the presence of cognitive impairment ranging from MCI to more severe impairment^23,24^. We used the MoCA as the primary measure for MCI, as it is the only measure of cognitive function that was administered by trained experimenters in both the FDNY and GRC responder cohorts. A conservative cut-off was applied to total MoCA scores to operationalize MCI (18 < MoCA < 23) because of the high reported sensitivity and specificity for MCI in a heterogeneous population (e.g., specificity was 91.3% in a meta-analysis)^25^. Prior work has also noted that this operational definition of MCI is highly accurate for identifying the presence of cortical atrophy in WTC-exposed populations (out of sample AUC=0.90)^26^ and when relying on biological definitions for ADRD^27^. However, other work suggests the MoCA is insensitive to localized cortical atrophy identified solely in the entorhinal cortex and fusiform gyri^28^. Therefore, to provide a more liberal estimate of prevalence based on the standard criteria outlined in the MoCA codebook, a less stringent cut-off of 17 < MoCA < 26 was used as a secondary operational definition of MCI in both WTC-exposed responder cohorts.

#### Domain-Specific Measures of Cognition

To allow for additional algorithmic routines to diagnose MCI, to facilitate comparison to meta-analytic estimates, to assess potential heterogeneity across diagnostic routines, and to assess the severity of impairments in specific domains, FDNY responders also completed an extensive battery of domain-specific cognitive assessments. The *Hopkins Verbal Learning Test (HVLT)* is a standard norm-referenced measure of verbal learning and verbal memory, which includes four subtests: total recall, delayed recall, retention, and recognition. The HVLT has sound psychometric properties, including high concurrent validity, inter-form reliability, and retest reliability^29,30^.

The *Trail Making Test (TMT-A & -B)* is another common norm-referenced measure of psychomotor speed and choice reaction speed that asks participants to draw a line as quickly as possible to connect a series of numbered circles that are miscellaneously scattered across a piece of paper (part A), and again alternating between circles with numbers and letters (part B). The construct validity of the TMT is supported by past studies^31^.

Having been described as a measure of cerebral dysfunction^32^, the *Symbol Digit Modalities Test (SDMT)* is a norm-referenced, paper-and-pencil, measure of processing and psychomotor speed that requires participants to replace abstract symbols with numbers using a reference key^32^. Due to high reliability, predictive validity, sensitivity, and specificity, the SDMT is a commonly used measure of processing speed in studies of multiple sclerosis^33^.

The *Controlled Oral Word Association (COWA)* test is a brief norm-referenced measure of verbal fluency that consists of three conditions, where participants are asked to say as many words as they can in one minute that begin with a given letter (F, A, or S). Test reliability and validity for the *COWA* are high^34^.

*The Boston Naming Test (BNT-30)* was administered to measure functional impairments related to loss of object recognition and verbal recall. This test asks subjects to name images of common objects and is designed to detect severe functional impairments related to visual memory and recall in patients with Alzheimer’s Disease^35^.

Finally, to assess individual differences in general academic aptitude and achievement, the *Wide Range Achievement Test (WRAT-Reading)* was administered. This brief measure of reading ability has been shown to exhibit strong retest reliability and no practice effects^36^.

#### Computerized Assessments of Cognition

We also fielded the CogState™ battery, a proprietary computerized program that tests for multiple domains of cognition and is reported to be sensitive to detecting dementia in aging populations^37,38^. The assessment consists of the Groton Maze Learning Test (GMLT), Detection Test (DET), Identification Test (IDN), One Card Learning Test (OCL), Continuous Paired Associate Learning Test (CPAL), and GMLT-Delayed. From these assessments, we calculated episodic memory, working memory, reaction speed, processing speed, cognitive throughput, visuospatial learning and recall.

#### Limitations Due to Cognitive Impairment

Subjective concern about changes in cognition and impact on daily functioning was measured using the *Cognitive Function Instrument (CFI)*. This self-report measure is often used to detect cognitive decline that results from aging in community samples^39^.

#### Possible Major Neurocognitive Disorder (NCD)

NIA-AA guidelines for the diagnosis of all-cause dementia^40^ were used to operationalize the presence of severe cognitive impairment. Specifically, FDNY responders were diagnosed with severe cognitive impairment if they met the following criteria: (1) impairment in one or more domain of verbal learning or memory as measured by the four subtests of the HVLT, indicated by an “impaired”, “moderate”, “severe” or “profound” level of impairment based on age-adjusted t-scores, (2) impairment in one or more test of executive function based on age-adjusted norms, measured by the TMT-A, TMT-B, SDMT, or COWA, (3) the presence of probable functional limitations, as indicated by three of more errors on the BNT-30 or self-reports of functional limitations due to cognitive impairment, as measured by a CFI score ≥ 1, and (4) average or above average performance on the WRAT-Reading to exclude poor cognitive test performance due to low academic achievement; Poor cognitive test performance due to traumatic brain injury or other known neurological disorders were excluded by study design.

### Objective Measures of Physical Functioning

*Short Physical Performance Battery (SPPB)* is a tool that consists of three tests used to evaluate lower extremity functioning, including balance, strength, and gait. Participants are asked by experimenters to demonstrate standing balance in three positions, lower limb strength by getting up and down from a chair, and walking at a normal speed^41^. *Handgrip Strength* is commonly employed in aging studies to measure frailty with high retest reliability and concurrent validity ^42^. For this assessment, participants are instructed to squeeze a hand-held dynamometer to determine maximum grip strength.

### Data Analytic Procedures

Analyses were conducted in R Studio Version (2023.12.0+369)^43^ using the following packages: ‘epiR’^44^, ‘lmtests’^45^, ‘sandwich’^46^, ‘effsize’^46^, ‘ggplot2’^47^, ’patchwork’^48^, ‘viridis’^49^. First, after calculating descriptive statistics to summarize sample characteristics, distributions of MoCA total scores were compared using histograms and density plots, and crude prevalences of MCI were estimated in FDNY and GRC responders using standard and conservative MoCA criteria (17 < MoCA < 26 and MoCA < 23, respectively). The prevalence of MCI was also estimated in FDNY responders using different algorithmic routines applied to domain specific measures of cognitive function, yielding multiple operational definitions of MCI, including Jak/Bondi criteria^50^, Petersen criteria^51^, and NIA-AA criteria^51^. To help visualize heterogeneity, determine clinical significance, and contextualize the magnitude of estimates, forest plots were used to display crude prevalence rates of MCI for different cohorts of WTC responders juxtaposed with meta-analytic estimates from different global, community, and clinical populations.

Second, generalized linear models were used to evaluate cohort differences (FDNY vs. GRC) in the prevalence of MCI, before and after controlling for age, sex, self-reported race/ethnicity, level of education, and county of residence. Specifically, robust Poisson regressions using a Huber-White sandwich estimator for variance estimation were used because odds ratios derived from logistic regressions overestimate relative risk ratios when outcomes are common (e.g., >10%)^52^. Similar Poisson regressions were then used to test whether FDNY responders and New York City Police Department (NYPD) officers, a subset of the GRC henceforth called GRC-NYPD, differ in the prevalence of MCI, as members of the FDNY and GRC-NYPD are similar regarding screening criteria for employment (both must complete a written examination and pass medical, social, and psychological tests) but differ in terms of their work experiences and exposures. Finally, similar generalized linear models were then estimated after excluding responders 65 years and older.

Third, a series of multiple linear regressions with heteroskedasticity-consistent standard errors (identical to Stata’s ‘robust’ option) were used to investigate the associations of domain-specific cognitive, psychological, and physical functional tests with mild cognitive impairment, defined as a binary variable (0=not impaired, 1=impaired) using conservative MoCA criteria (total scores < 23) and adjusting for age-related differences in test performance. Finally, raw scores from norm-referenced tests were transformed to age-adjusted scaled scores (either t-scores or z-scores), accompanied by their recommended interpretations (e.g., “average”, “low average”, “borderline”, “impaired”, “moderate”, “severe”, and “profound”) to help determine the severity of impairments across specific domains of cognitive function.

## Results

Sample characteristics of WTC-exposed FDNY and GRC responders are reported in Table 1 and Table S1. Descriptive statistics for cognitive, psychological, and physical function measures of FDNY responders are summarized in Table S2.

**Table 1.** Sample Characteristics.

|  |  | WTC-Exposed<br>FDNY Responders<br>(*n* = 343) |  | WTC-Exposed<br>GRC Responders<br>(*n* = 7102) |  |
| --- | --- | --- | --- | --- | --- |
|  |  | *M/f* | *SD/%* | *M/f* | *SD/%* |
| **Age** |  | 59.58 | 6.25 | 57.39 | 6.31 |
|  | Over 65 Years | 88 | 25.66 | 917 | 12.91 |
|  | Under 65 Years | 255 | 74.34 | 6185 | 87.09 |
| **Sex** |  |  |  |  |  |
|  | Male | 338 | 98.54 | 6443 | 90.72 |
|  | Female | 5 | 1.46 | 659 | 9.28 |
| **Race/Ethnicity** |  |  |  |  |  |
|  | White | 322 | 93.88 | 5879 | 82.78 |
|  | Black | 7 | 2.03 | 812 | 11.43 |
|  | Latino | 7 | 2.03 | 314 | 4.42 |
|  | Asian | 0 | 0.00 | 43 | 0.61 |
|  | Other | 7 | 2.03 | 54 | 0.76 |
| **Education** |  |  |  |  |  |
|  | No High School Diploma | 0 | 0.00 | 1185 | 16.69 |
|  | High School Diploma | 37 | 10.79 | 1006 | 14.17 |
|  | Some College/Tech. School | 177 | 51.60 | 2993 | 42.14 |
|  | Bachelor's Degree | 102 | 29.74 | 1522 | 21.43 |
|  | Graduate School or Higher | 27 | 7.87 | 396 | 5.58 |
| **Occupation on 9/11/2001** |  |  |  |  |  |
|  | Civilian Public Sector | 0 | 0.00 | 341 | 4.80 |
|  | Construction Worker | 0 | 0.00 | 583 | 8.21 |
|  | Emergency Medical Services (EMS) | 27 | 7.87 | 0 | 0.00 |
|  | EMS – Supervisor | 1 | 0.29 | 0 | 0.00 |
|  | Firefighter | 308 | 89.80 | 0 | 0.00 |
|  | Firefighter – Supervisor | 7 | 2.03 | 0 | 0.00 |
|  | Law Enforcement | 0 | 0.00 | 3713 | 52.28 |
|  | Unemployed | 0 | 0.00 | 10 | 0.14 |
|  | Other | 0 | 0.00 | 2331 | 32.82 |
|  | Missing | 0 | 0.00 | 124 | 1.75 |
| **County of Residence** |  |  |  |  |  |
|  | Suffolk | 182 | 53.06 | 3416 | 44.26 |
|  | Nassau | 155 | 45.19 | 3331 | 42.47 |
|  | Other | 6 | 1.74 | 943 | 13.28 |

**Notes.** *n* = sample size. *f* = frequency. *M* = mean. *SD* = standard deviation. % = percent

### Prevalence of Mild Cognitive Impairment (MCI)

The prevalence of MCI determined using different diagnostic criteria for the FDNY responders, and using conservative and standard cut-off criteria applied to MoCA total scores for both FDNY and GRC responders are reported in Table 2. In the FDNY cohort, different operational definitions of MCI (Jak/Bondi, Petersen, NIA-AA) yielded similar prevalence estimates for the full sample (range= 51.91% to 58.11%) and for FDNY responders under 65 years old (range=52.57% to 60.32%). When MCI was determined using standard (18 - 25) and conservative (< 23) criteria applied to total MoCA scores, prevalence estimates for FDNY responders were higher (71.72%) and lower (24.78%) than prevalence estimates based on canonical operational definitions of MCI. To contextualize the clinical significance of these findings, crude prevalence estimates of MCI for WTC-exposed cohorts based on different operational definitions are descriptively juxtaposed with meta-analytic estimates of MCI in Figure 1. Notably, crude prevalence rates were visually higher for WTC responders from both the FDNY and GRC cohorts compared to meta-analytic estimates from community and clinical populations.

**Figure 1.**
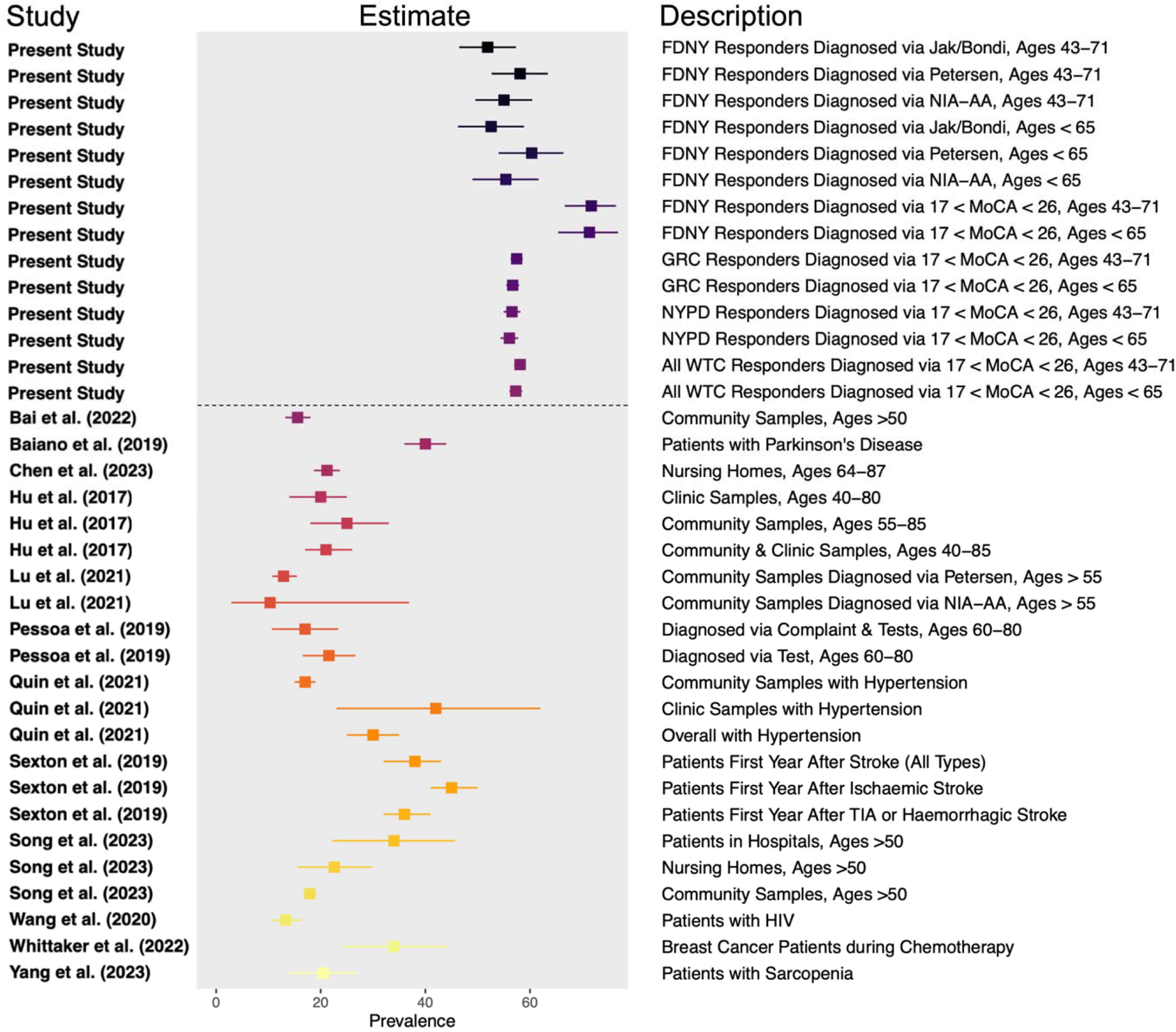
Prevalence of Mild Cognitive Impairment in WTC Responders and from Meta-Analyses of Community and Clinical Populations **Notes.** Squares mark the number of estimated cases of MCI per 100 individuals at risk with horizontal lines denoting 95% confidence intervals.

**Table 2.** Prevalence of Mild and Severe Cognitive Impairment among WTC-Exposed Responders.

| Cohort | Diagnostic Criteria | *n* | Cases | Crude Period Prevalence | Lower 95% | Upper 95% |
| --- | --- | --- | --- | --- | --- | --- |
| FDNY Responders |  |  |  |  |  |  |
| Mild Cognitive Impairment | Jak/Bondi | 341 | 177 | 51.91 | 46.46 | 57.32 |
|  | Petersen | 339 | 197 | 58.11 | 52.66 | 63.42 |
|  | NIA-AA | 338 | 186 | 55.03 | 49.55 | 60.42 |
|  | 17 < MoCA < 26 | 343 | 246 | 71.72 | 66.63 | 76.43 |
|  | 18 < MoCA < 23 | 343 | 85 | 24.78 | 20.30 | 29.70 |
| Severe Cognitive Impairment | NIA-AA | 343 | 17 | 4.96 | 2.91 | 7.82 |
|  | MoCA < 19 | 343 | 7 | 2.04 | 0.82 | 4.16 |
| FDNY Responders under 65 years old |  |  |  |  |  |  |
| Mild Cognitive Impairment | Jak/Bondi | 253 | 133 | 52.57 | 46.22 | 58.86 |
|  | Petersen | 252 | 152 | 60.32 | 53.99 | 66.40 |
|  | NIA-AA | 251 | 139 | 55.38 | 49.00 | 61.63 |
|  | 17 < MoCA < 26 | 255 | 182 | 71.37 | 65.40 | 76.84 |
|  | 18 < MoCA < 23 | 255 | 62 | 24.31 | 19.18 | 30.06 |
| Severe Cognitive Impairment | NIA-AA | 255 | 11 | 4.31 | 2.17 | 7.59 |
|  | MoCA < 19 | 196 | 5 | 1.96 | 0.64 | 4.52 |
| GRC Responders |  |  |  |  |  |  |
| Mild Cognitive Impairment | 17 < MoCA < 26 | 7102 | 4081 | 57.46 | 56.30 | 58.62 |
|  | 18 < MoCA < 23 | 7102 | 1286 | 18.11 | 17.22 | 19.02 |
| Severe Cognitive Impairment | MoCA < 19 | 7102 | 230 | 3.24 | 2.84 | 3.68 |
| GRC Responders under 65 years old |  |  |  |  |  |  |
| Mild Cognitive Impairment | 17 < MoCA < 26 | 6185 | 3507 | 56.70 | 55.46 | 57.94 |
|  | 18 < MoCA < 23 | 6185 | 1064 | 17.20 | 16.27 | 18.17 |
| Severe Cognitive Impairment | MoCA < 19 | 6185 | 178 | 2.88 | 2.48 | 3.33 |
| GRC NYPD Responders |  |  |  |  |  |  |
| Mild Cognitive Impairment | 17 < MoCA < 26 | 3713 | 2100 | 56.56 | 54.95 | 58.16 |
|  | 18 < MoCA < 23 | 3713 | 625 | 16.83 | 15.64 | 18.08 |
| Severe Cognitive Impairment | MoCA < 19 | 3713 | 97 | 2.61 | 2.12 | 3.18 |
| GRC NYPD Responders under 65 years old |  |  |  |  |  |  |
| Mild Cognitive Impairment | 17 < MoCA < 26 | 3412 | 1913 | 56.07 | 54.38 | 57.74 |
|  | 18 < MoCA < 23 | 3412 | 548 | 16.06 | 14.84 | 17.34 |
| Severe Cognitive Impairment | MoCA < 19 | 3412 | 82 | 2.40 | 1.92 | 2.97 |

**Notes.** Crude period prevalence is the number of cases per 100 individuals at risk. *n* = sample size. Lower and Upper 95% = bounds of confidence interval.

### Cohort Differences in MCI: FDNY vs. GRC

On average, MoCA total scores were significantly lower for FDNY responders (mean = 23.83, median = 24) compared to GRC responders (mean = 24.51, median = 25), as indicated by Welch’s t-test (*p* < .001) and Wilcoxon rank sum test with continuity correction (*p* < .001), amounting to a small standardized mean difference (Cohen’s *d* with Hedge’s correction = .24, 95% CI = [.13, .35]). The crude prevalence of MCI was also higher for FDNY than GRC responders when using standard MoCA criteria (71.72% vs. 57.46%) and when using conservative MoCA criteria (24.78% vs. 18.11%). The observed distributions of MoCA total scores for FDNY and GRC responders that were used to determine MCI are depicted in Figure 2.

**Figure 2.**
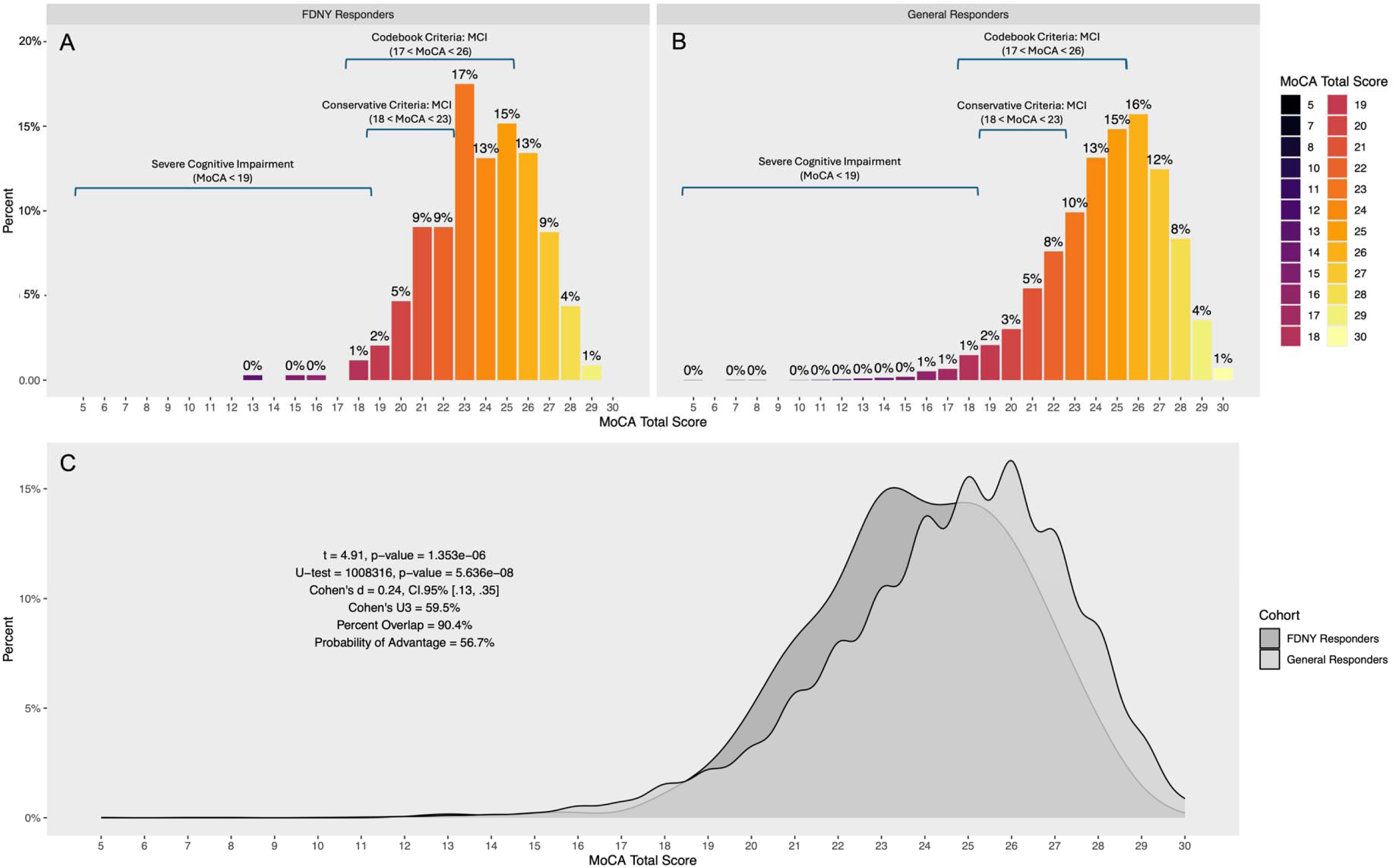
Comparing Distributions of Cognitive Impairment Measured by the MoCA in Two Cohorts of WTC Responders **Notes.** Level of cognitive impairment was measured using MoCA total scores, and MCI was determined using codebook criteria and conservative criteria. “0%” indicates less than 0.50%. To help ease interpretation of the statistics reported in panel C, the standardized mean difference between cohorts is 0.24, approximately 59.5% of responders from the general responder cohort have MoCA total scores above the mean of FDNY responders, 90.4% of scores from the two cohorts overlap, and there is a 56.7% chance that a WTC responder picked at random from the GRC will have better cognitive performance on the MoCA than a WTC responder picked at random from the FDNY cohort. Crude prevalence rates based on the depicted criteria are reported in Table 2.

Results of Poisson regressions testing cohort differences in MCI are reported in Table 3. When based on conservative MoCA criteria, MCI prevalence remained significantly higher for WTC-exposed FDNY responders, compared to WTC-exposed responders from the GRC, adjusting for differences in sample characteristics (adjusted *RR*=1.53, 95% CI = [1.24, 1.88], *p* < .001), and similarly, when compared to GRC-NYPD law enforcement responders (adjusted *RR*=1.43, 95% CI = [1.14, 1.80], *p* = .002). Reported in Table 3, the results of null hypothesis significance tests remained unchanged when prevalence rates were based on standard MoCA criteria, when comparing FDNY responders to GRC-NYPD law enforcement responders, and after excluding responders 65 years and older.

**Table 3.**
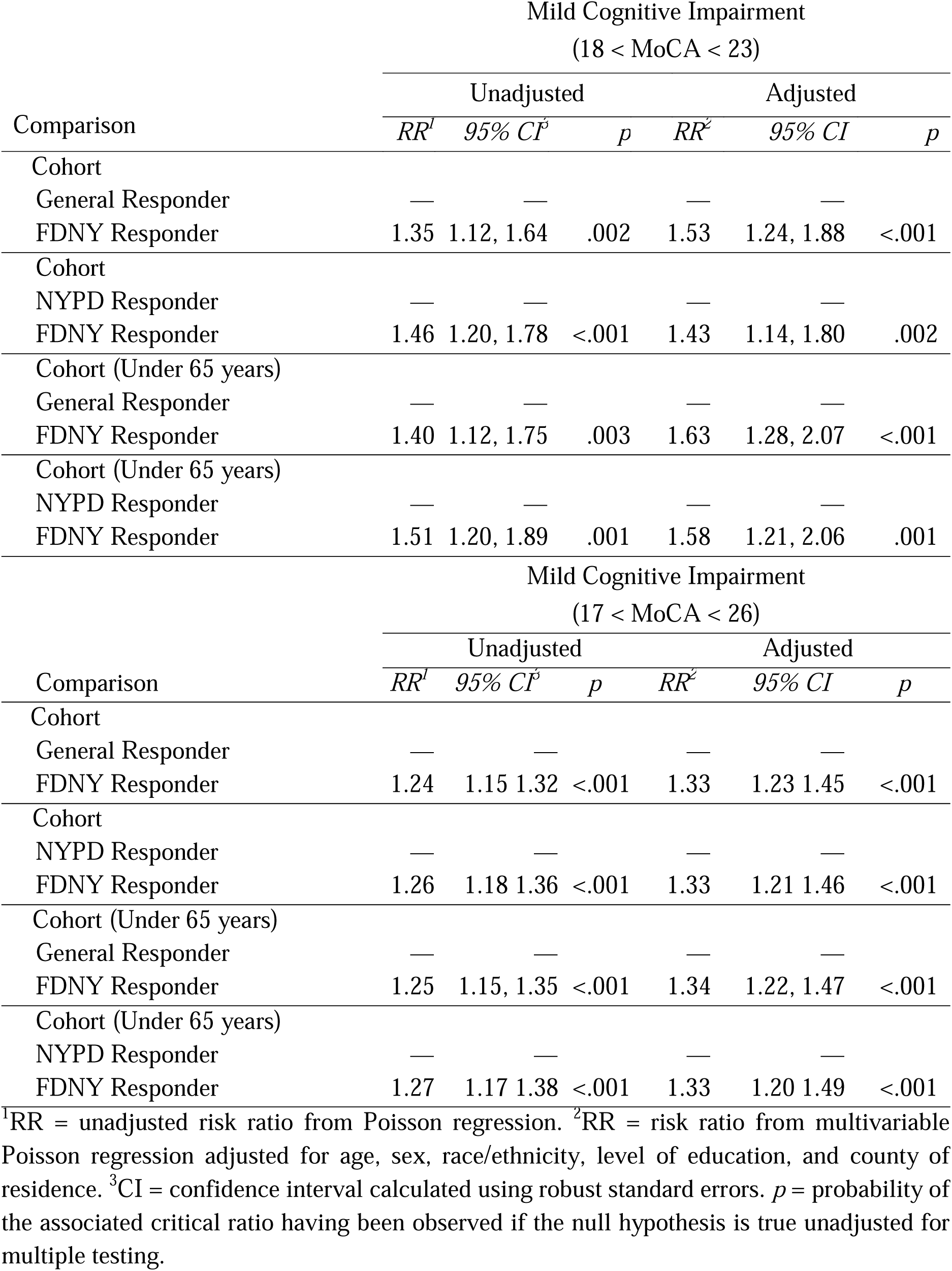
Results of Poisson Regressions Testing Cohort Differences in Prevalence Rates of Cognitive Impairment Measured Using the Montreal Cognitive Assessment (MoCA)

### Domain-Specific Correlates of MCI

To examine the domain-specific features of cognitively impaired FDNY responders, Figure 3 displays the age-adjusted correlates of mild-to-severe cognitive impairment diagnosed using MoCA total scores < 23. Specifically, standardized regression coefficients (β) from a series of multivariable linear regressions are plotted with 95% confidence intervals and p-values calculated using heteroskedasticity-consistent standard errors. These coefficients quantify whether responders with MCI, on average, exhibit deficits in a specific domain of cognitive function, while adjusting for the effects of age. Compared to unimpaired responders, responders with MCI exhibited deficits in all experimenter-administered tests of cognitive function, including total recall (β = -0.65, *p* < .001), delayed recall (β = -0.59, *p* < .001), retention (β = -0.29, *p* = .038), and recognition (β = -0.49, *p* < .001) measured by the HVLT, visual memory measured by the BNT (β = -0.39, *p* = .016), reading comprehension measured by the WRAT-4 Reading (β = -0.41, *p* = .002), psychomotor speed measured by the TMT-A (β = -0.49, *p* < .001), choice reaction speed and attentional shifting measured by the TMT-B (β = -0.65, *p* < .001), processing speed measured by the SDMT (β = -0.34, *p* = .004), and verbal fluency measured by the COWA (β = -0.73, *p* = .002).

**Figure 3.**
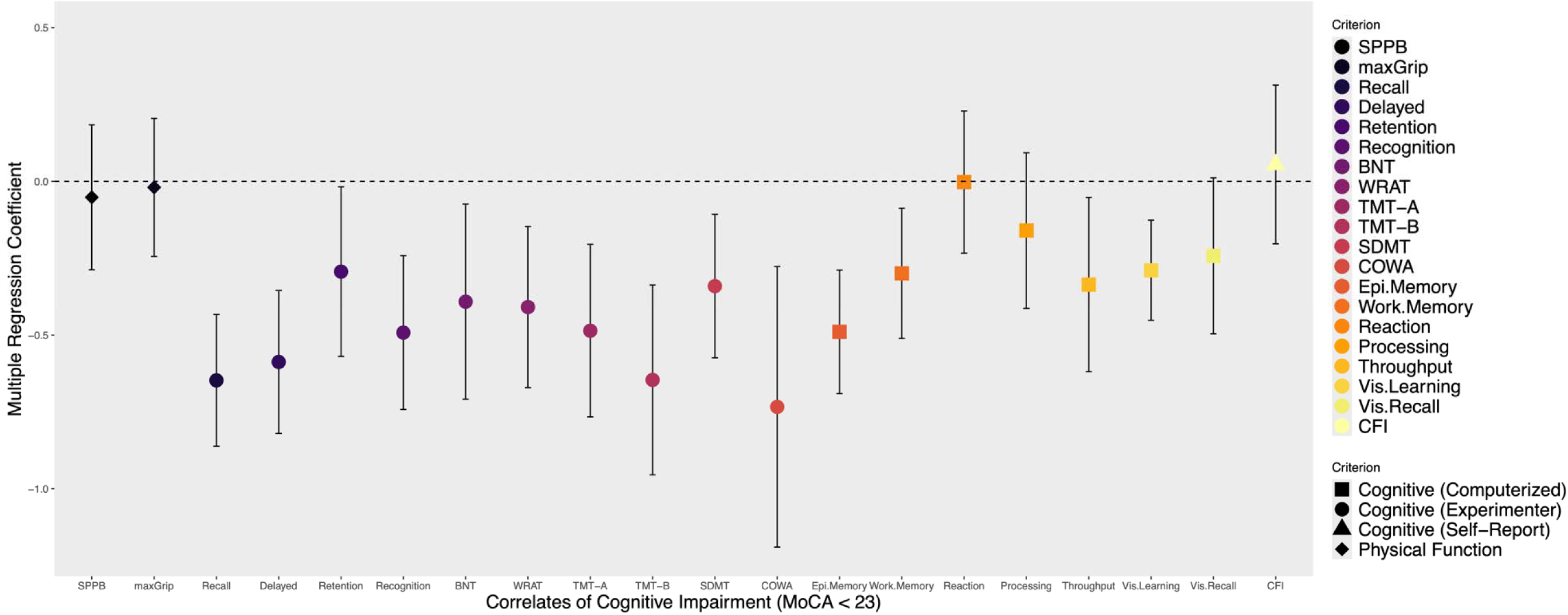
Domain-Specific Correlates of Mild-to-Severe Cognitive Impairment in WTC-Exposed FDNY Responders **Notes.** Colored shapes denote multiple regression coefficients for mild cognitive impairment (coded 0 = unimpaired, 1 = impaired) and domain-specific continuous measures of cognitive and physical function (reported on the x-axis) standardized with respect to the criterion (STDY) and adjusted for the effects of age. Vertical bars donote 95% confidence intervals calculated using heteroskedasticity-consistent standard errors. Coefficients with 95% confidence intervals that do not include zero (i.e., vertical bars that do not cross the dashed horizontal line) are statistically significant at *p* < .05. Scores for TMT-A and TMT-B were reserve coded so lower scores indicate worse performance.

On average, compared to unimpaired responders, responders with MCI also exhibited deficits in computerized assessments of episodic memory (β = -0.49, *p* < .001), working memory (β = -0.30, *p* = .006), cognitive throughput (β = -0.34, *p* = .021), and visual learning (β = -0.29, *p* < .001). Notably, self-reports of subjective cognitive concerns (average CFI scores) were not significantly different when comparing cognitively impaired and unimpaired responders (β = 0.06, *p* = .679). Similarly, indicators of physical function were not significantly different when comparing cognitively impaired and unimpaired responders, including lower extremity function measured by the SPPB (β = -0.05, *p* = .666) and maximum grip strength (β = -0.02, *p* = .864).

### Prevalence of Severe Impairments and Domain-Specific Impairments

The severity of impairments across specific domains of cognition that were measured by norm-referenced tests are depicted in Figure 4 and reported in Table S3. The age-adjusted prevalence rates of moderate (0.58% - 3.50%), severe (1.17% - 6.12%), and profound (8.75% - 10.79%) impairments were high for tests of verbal learning and memory (measured by the HVLT; panels A-D), compared to certain executive functions, specifically verbal fluency and processing speed (measured by the COWA & SDMT; panels E & H), whereby moderate impairments were quite rare (0% - 0.29%) and severe and profound impairments were entirely absent. On the other hand, the age-adjusted prevalence rates of moderate (0.87% - 1.46%), severe (0% - 0.29%), and profound (2.62% - 9.62%) impairments were more common for other executive functions, like psychomotor speed and choice reaction speed (measured by the TMT-A & TMT-B). Finally, following NIA-AA diagnostic guidelines for early-onset dementia, 4.96% of WTC-exposed FDNY responders met the criteria for severe cognitive impairment (95% CI = [2.91% to 7.82%]). Furthermore, this prevalence rate remained largely unchanged after excluding FDNY responders over the age of 65 years (Table 2).

**Figure 4.**
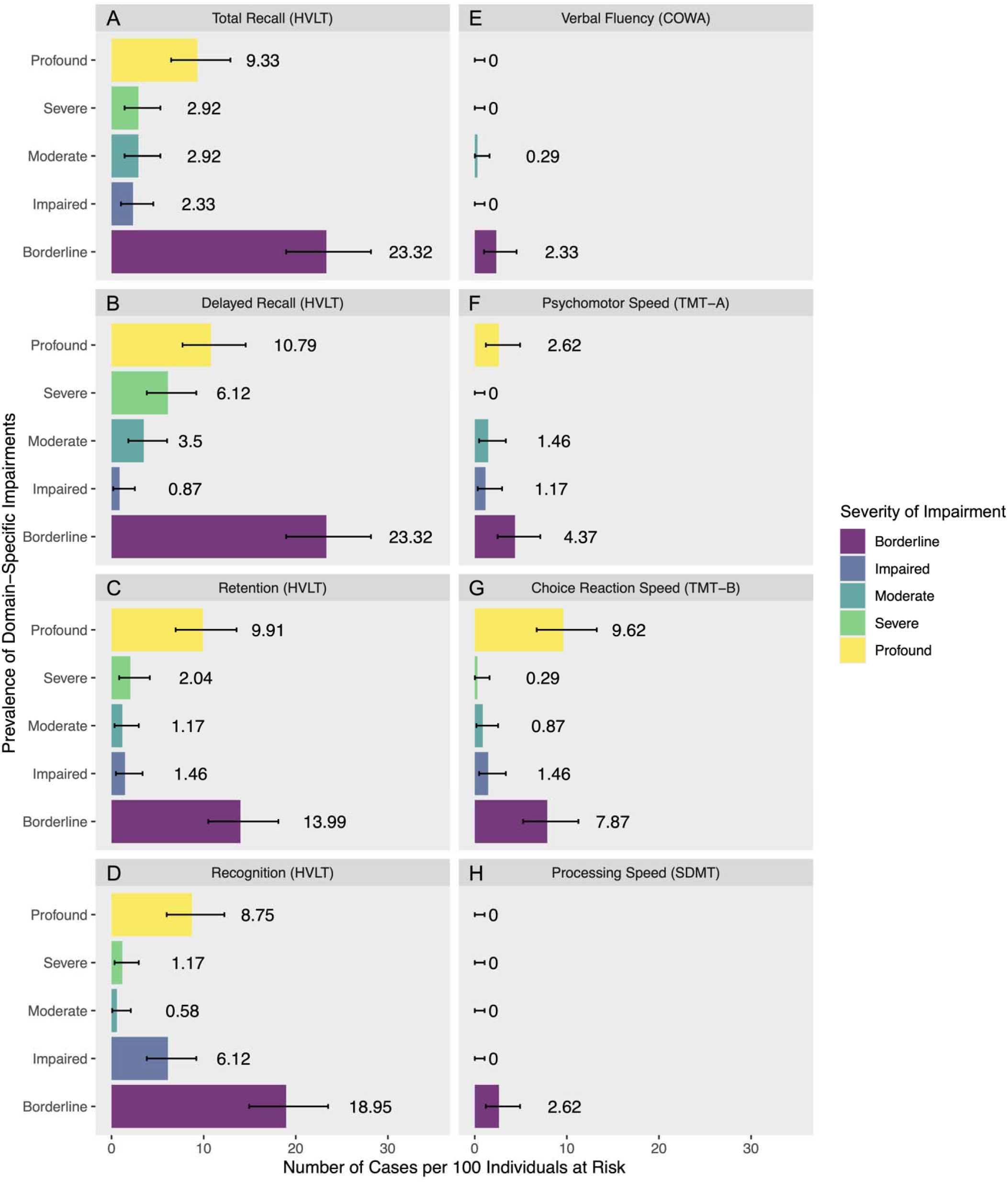
Severity of Domain-Specific Cognitive Impairments in WTC-Exposed FDNY Responders **Notes.** Prevalence rates of domain-specific impairments are plotted across levels of severity based on age-adjusted norms. Horizontal bars donote 95% confidence intervals calculated using the exact method.

## Discussion

The present study is the first to investigate cognitive impairment in WTC-exposed-FDNY-responders using objective and well-validated tests of global and domain-specific neurocognitive function. This fills a central gap in information about FDNY responders, as to date, data collection efforts have used state-of-the-art methodologies to collect information only for GRC WTC responders who participate in the Stony Brook WTC Health and Wellness Program. Because WTC exposure may differ between responder cohorts, it was important to document the prevalence of mild and severe cognitive impairment in the FDNY cohort as well. We found that the unadjusted prevalence of MCI among FDNY responders varied from approximately 26% to 72% depending on the operational definition of MCI, and prevalence rates were significantly higher than GRC responders in adjusted models. Compared with that previously reported in the general population, as well as different clinical populations, the prevalence of MCI is noticeably higher for WTC responders, including both responders from the FDNY and GRC. Clinical populations included residents of nursing homes^15,16^, patients in hospitals^15^, and patients with Parkinson’s disease^17^, clinical hypertension^18^, scarpenia^19^, ischemic and hemorrhagic stroke^20^, HIV^52^, and breast cancer during chemotherapy^21^. These stark comparisons indicate that occupational and environmental exposures should be considered more carefully in future studies and prevention efforts as potential pathogenic exposures for cognitive impairment. The prevalence of MCI among FDNY responders also appeared higher than meta-analytic estimates based on different diagnostic criteria, including Petersen and NIA-AA criteria, and irrespective of whether MCI was diagnosed based on subjective complaints or objective tests. Moreover, prevalence estimates of MCI remained largely unchanged after excluding responders 65 years and older, suggesting that such high prevalence rates among WTC responders are not attributable to common geriatric diseases.

Results from an extensive battery of domain-specific neurocognitive tests indicated that cognitively impaired FDNY responders suffer broadly across cognitive domains from deficits in verbal learning, verbal memory, processing speed, and choice reaction speed. Moreover, these deficits were observed after adjusting for potential age-related differences in neurocognitive function. Among the specific domains that were measured in the present study, approximately 10% to 16% of WTC-exposed-FDNY-responders exhibited severe-to-profound deficits in verbal learning and memory, and approximately 3% to 10% exhibited severe-to-profound deficits in psychomotor speed and choice reaction speed. Moreover, based on NIA-AA diagnostic guidelines, approximately 5% of WTC-exposed-FDNY-responders exhibited impairment in multiple domains of cognition and probable functional limitations, despite average or above average academic aptitude, supporting the diagnosis of severe cognitive impairment. These findings underscore the need for clinicians to anticipate mild and severe cognitive impairment characterized by diffuse impairments across multiple domains when screening patients and highlight treatment planning for trauma-exposed occupational cohorts as a high priority for public health efforts moving forward.

To date, there is limited research on MCI and major NCD in occupational cohorts, and little is known about the potential impacts of occupational or environmental exposures on cognitive impairment and underlying neurodegenerative disease. The biomarkers of severe neurodegenerative diseases have been detected in forensic autopsies of children, adolescents, and young adults exposed to fine particulate matter, combustion and friction ultrafine particulate matter, and industrial nanoparticles (NPs), including biomarkers of Alzheimer’s disease, Parkinson’s disease, frontotemporal lobar degeneration, and amyotrophic lateral sclerosis (ALS)^53^. These biomarkers include neuronal cytoplasmic tau in the substantia nigra, tau positive neurites in the brainstem, intracytoplasmic tau and positive tau nuclei, and b-amyloid plaques in the temporal cortex^53^. These findings are consistent with structural MRI studies that have found links between exposure to air pollutants, cortical atrophy, and disturbances in white matter connections^54,55^, as well as findings from animal models, which show that increases in amyloid-β40 and amyloid-β42 can be caused by prolonged exposure to air pollution^56^. Inhalation of air pollutants has also been associated with an increased risk of MCI *via* a process of tauopathy in police and other emergency personnel who responded to the WTC attacks^57^, and that impairment has been linked to cortical atrophy^28^. Using National Death Index data, however, FDNY WTC responders were not found to have an increased incidence of ALS^58^, but additional studies are needed to examine how occupational, environmental and disaster-related exposures could influence neuropathology in this and other cohorts.

Prior studies have been limited by a focus on using short-form and computerized cognitive batteries for assessing cognition and for diagnosing MCI in general responders. While valid, those studies could not compare the prevalence of MCI using different diagnostic criteria as is reported here, and they could neither specifically target which domains of cognition were impacted by MCI nor diagnose severe impairments using a large battery of norm-referenced tests. Additionally, this study clarifies that while verbal learning and verbal memory, psychomotor speed, and choice reaction speed are the most strongly impacted domains of neurocognitive function, episodic memory, working memory, and visual-spatial function are also impaired in responders with MCI. This diffuse pattern of impairment across cognitive domains is not consistent with MCI or dementia due to Alzheimer’s Disease, which tends to cause drastic reductions in episodic memory prior to causing declines in other domains of cognition.

### Strengths & Limitations

A strength of our study was the administration of an extensive battery of objective neurocognitive assessments, including tests of global and domain-specific neurocognitive function. This enabled the diagnosis of MCI using different algorithmically determined operational definitions in WTC-exposed FDNY responders, while simultaneously enabling a thorough domain-specific characterization of neurocognitive deficits among impaired responders. Another strength was our ability to directly compare differently exposed WTC groups – FDNY vs. GRC and FDNY vs. GRC-NYPD. A third strength was finding similar MCI prevalence rates across secondary operational definitions of MCI including Jak/Bondi criteria, Petersen criteria, and NIA-AA criteria, while estimates of MCI prevalence were even higher when using traditional MoCA criteria but noticeably lower when using more conservative MoCA criteria, as employed in past studies of MCI in GRC responders^2^.

There are several limitations. First, the present study was cross-sectional and, consequently, provides no information about the rate of change in cognitive decline, potential reversion of MCI over time, long-term incidence, or prognosis. Related, despite following NIA-AA guidelines for the diagnosis of severe cognitive impairment^40^, the lack of repeated measures precludes determination of the “continuing decline” criteria for major NCD. Second, although randomly selected, participants voluntarily chose whether to enroll in the study, introducing the potential for self-selection bias. Third, comparisons to non-WTC responders were descriptive due to potential comparability issues regarding operational definitions of MCI and differences in statistical methodologies. Fourth, apart from the MoCA, GRC-responders did not complete experimenter administered neurocognitive assessments and, therefore, could not be diagnosed using a wide variety of well-established algorithmic routines like FDNY-responders, limiting the number of directly comparable estimates across the FDNY and GRC. However, the MoCA was administered to both FDNY and GRC responders, enabling comparison of prevalence estimates of MCI across these cohorts. Fifth, like all non-experimental studies, cause-effect relationships cannot be discerned. However, such high prevalence rates of MCI among WTC-exposed responders, even higher than clinical samples of patients with serious neurological and medical conditions, is highly suggestive of a causal effect of the occupational exposures endured during the response efforts to the WTC attacks.

Similarly, such high prevalence rates of severe-to-profound impairments across specific domains of cognition (∼3% to 16%), as well as a high prevalence rate of severe impairments and functional limitations indicative of a possible major NCD (∼5%) are unlikely to be explained by common geriatric disorders, as these prevalence rates were considerably higher than what would be expected in the general population under 65 years of age (e.g., 0.01% to 0.50% prevalence for early onset dementia)^16–20^.

Nevertheless, future analyses will investigate the relationship between level of WTC exposures and cognitive dysfunction. Sixth, no genetic testing was done among WTC-exposed FDNY responders, so we cannot rule out the possibility that APoE e4 or other genetic factors might contribute to the high rates of cognitive impairment observed in the present study. However, this seems unlikely as previous research in GRC responders has shown the link between WTC exposure severity and cumulative hazard of cognitive impairment is *not* explained by polygenic risk for Alzheimer’s Disease^59^. Seventh, FDNY responders were predominately White and male which is reflected in this study’s source population. Consequently, the generalizability of findings to more diverse populations might be limited. Finally, though cognitive symptoms are one of the most common functional indicators of neurodegenerative disease, the present study did not validate findings using neuroimaging or other biomarkers. Future work should seek to corroborate the present findings, not only using longitudinal studies of cognition, but also using targeted neuroimaging and serological studies.

### Conclusion

In the first study to examine the prevalence of MCI and possible major NCD in WTC-exposed-FDNY-responders, we identified a high level of MCI compared to WTC-exposed-GRC-responders in analyses that controlled for differences in sample characteristics and demographic factors. Crude prevalence rates of MCI were also higher than recent meta-analytic estimates from different community and clinical populations, and rates of severe cognitive impairment were higher than expected in the general population based on meta-analyses of early onset dementia. Cognitive symptoms were widespread, with deficits that spanned domains of cognitive function, including verbal learning, memory, psychomotor speed, and choice reaction speed. Such high rates of cognitive impairments are concerning. Researchers and clinicians focused on screening and prevention efforts should give occupational and environmental exposures serious consideration as potential pathogenic risk factors for cognitive impairment. Future research is needed to examine longitudinal rates of cognitive decline in FDNY-responders and other cohorts with significant occupational and environmental exposures, as well as the impact of comorbidities and treatments on rate of decline. Following occupational exposures, clinicians should include screening and treatment planning for mild-to-severe cognitive impairment, and this should be a high priority for public health efforts moving forward.

## Data Availability Policy and Statement

Data are not publicly available. Requests for deidentified data will be considered and will require approval from principal investigators and the IRB at the first author’s home institution.

## Statement of Competing Interests

The authors have no competing interests to report.

## Funding & Acknowledgements

The first author conducted analyses and drafted the manuscript. All authors contributed to participant recruitment and data collection, provided critical revisions to the manuscript and approved a final version. Data collection was funded by U01OH012258 awarded to S.A.P.C and C.B.H., R01AG049953 awarded to S.A.P.C., CDC 2011-200-39361 awarded to B.J.L., and by the SUNY Research Foundation. The first author was also awarded and partially funded by R21AG074705-01.

## Supplemental Materials

### Recruitment and Sample Characteristics

We recruited 322 participants via letters, of whom 13.66% (*n*=44) dropped out before screening due to a lack of interest, and 5.28% (*n*=17) were deemed to be ineligible due to the stated exclusion criteria. An additional 94 participants were recruited via follow-up calls, of whom 12 were deemed to be ineligible for the study. This resulted in an analytic sample of *n* = 343 participants recruited from letters (*n*=261, 76.09%) or follow-up calls (*n*=81, 23.91%). Sample characteristics are reported in Table 1, contrasted with those of the comparison cohort comprising WTC responders from the GRC. Briefly, the FDNY and GRC samples were predominately male and White, but there was more racial diversity among GRC responders than FDNY responders, and there were more female GRC responders than female FDNY responders. GRC responders were also slightly younger (mean = 57.30 years), on average, compared with the FDNY responders (mean = 59.58 years), but the cohorts had the same range of ages by design (41 to 71 years). There was considerable variation in educational attainment, but most had at least some college and approximately 30% had a university degree, while fewer GRC responders were as highly educated.

---

**Table S1.** Characteristics of Source Population for WTC-exposed FDNY Responders.

|  | Enrolled<br>from Letter | All Letters<br>Sent | Source Population |
| --- | --- | --- | --- |
| n | 343 | 2446 | 4198 |
| Mean CFI score (SD) | 1.52 (2.4) | 0.07 (0.2) | 1.48 (2.5) |
| Median CFI score (IQR) | 0.5 (0-2) | 0 (0-0) | 0 (0-2) |
| Age on 9/11 (SD) | 38.4 (6.3) | 37.6 (6.4) | 37.3 (6.7) |
| Race |  |  |  |
| White | 322 (94%) | 2307 (94%) | 3944 (94%) |
| Black | 7 (2%) | 56 (2%) | 103 (2%) |
| Hispanic | 7 (2%) | 69 (3%) | 128 (3%) |
| Other | 7 (2%) | 14 (1%) | 23 (1%) |
| Male | 338 (99%) | 2360 (96%) | 4029 (96%) |

**Table S2.**
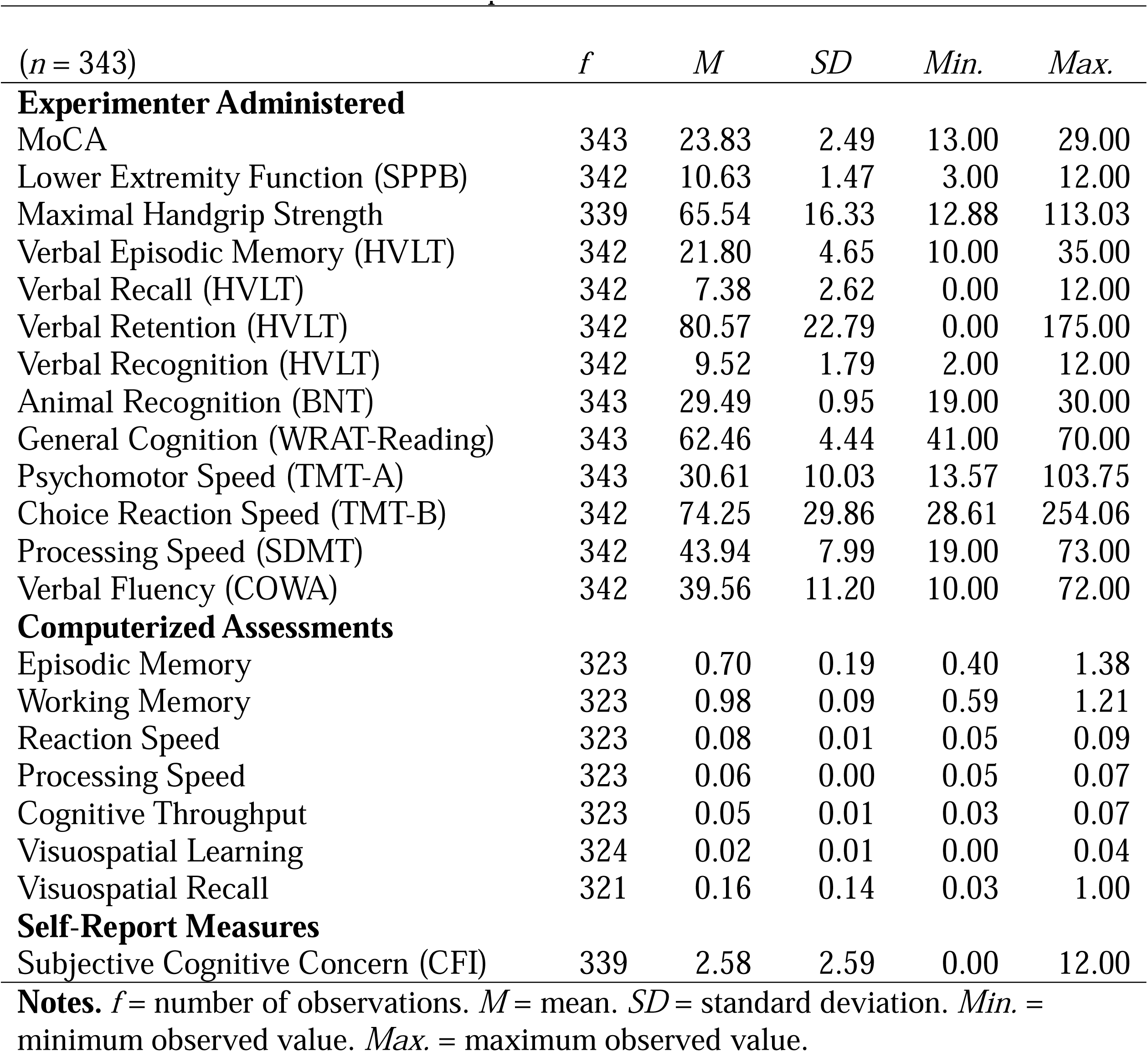
Descriptive Statistics for Neurocognitive, Psychiatric, and Physical.

**Table S3.** Prevalence of Domain-Specific Impairments by Level of Severity Among WTC-Exposed FDNY Responders.

| Domain (Test) | Severity | Period<br>Prevalence | Lower<br>95% | Upper<br>95% |
| --- | --- | --- | --- | --- |
| Total Recall (HVLТ) | Borderline | 23.32 | 18.95 | 28.16 |
|  | Impaired | 2.33 | 1.01 | 4.54 |
|  | Moderate | 2.92 | 1.41 | 5.30 |
|  | Severe | 2.92 | 1.41 | 5.30 |
|  | Profound | 9.33 | 6.47 | 12.91 |
| Delayed Recall (HVLТ) | Borderline | 23.32 | 18.95 | 28.16 |
|  | Impaired | 0.87 | 0.18 | 2.53 |
|  | Moderate | 3.50 | 1.82 | 6.03 |
|  | Severe | 6.12 | 3.83 | 9.21 |
|  | Profound | 10.79 | 7.71 | 14.56 |
| Retention (HVLТ) | Borderline | 13.99 | 10.50 | 18.12 |
|  | Impaired | 1.46 | 0.47 | 3.37 |
|  | Moderate | 1.17 | 0.32 | 2.96 |
|  | Severe | 2.04 | 0.82 | 4.16 |
|  | Profound | 9.91 | 6.96 | 13.58 |
| Recognition (HVLТ) | Borderline | 18.95 | 14.94 | 23.51 |
|  | Impaired | 6.12 | 3.83 | 9.21 |
|  | Moderate | 0.58 | 0.07 | 2.09 |
|  | Severe | 1.17 | 0.32 | 2.96 |
|  | Profound | 8.75 | 5.98 | 12.25 |
| Psychomotor Speed (TMT-A) | Borderline | 4.37 | 2.47 | 7.11 |
|  | Impaired | 1.17 | 0.32 | 2.96 |
|  | Moderate | 1.46 | 0.47 | 3.37 |
|  | Severe | 0.00 | 0.00 | 1.07 |
|  | Profound | 2.62 | 1.21 | 4.92 |
| Choice Reaction Speed (TMT-B) | Borderline | 7.87 | 5.25 | 11.25 |
|  | Impaired | 1.46 | 0.47 | 3.37 |
|  | Moderate | 0.87 | 0.18 | 2.53 |
|  | Severe | 0.29 | 0.01 | 1.61 |
|  | Profound | 9.62 | 6.72 | 13.25 |
| Verbal Fluency (COWA) | Borderline | 2.33 | 1.01 | 4.54 |
|  | Impaired | 0.00 | 0.00 | 1.07 |
|  | Moderate | 0.29 | 0.01 | 1.61 |
|  | Severe | 0.00 | 0.00 | 1.07 |
|  | Profound | 0.00 | 0.00 | 1.07 |
| Processing Speed (SDMT) | Borderline | 2.62 | 1.21 | 4.92 |
|  | Impaired | 0.00 | 0.00 | 1.07 |
|  | Moderate | 0.00 | 0.00 | 1.07 |
|  | Severe | 0.00 | 0.00 | 1.07 |
|  | Profound | 0.00 | 0.00 | 1.07 |

**Notes.** Period prevalence is the number of cases per 100 individuals at risk. Lower and Upper 95% = bounds of confidence interval.

## Notes

### Competing Interest Statement

The authors have declared no competing interest.

### Author Declarations

All protocols were approved by the Institutional Review Board at Stony Brook University (CORIHS IRB#2021-00295), and participants provided informed oral and written consent.

